## Supplementary data for "Plasma metabolomics profiling and machining learning-driven prediction of nonalcoholic steatohepatitis"

#### Metabolomics analysis methods

##### GC-MS/MS analysis

The GC-MS/MS analysis of FAs, OAs, and AAs was performed using a Shimadzu 2010 plus gas chromatograph interfaced with a Shimadzu TQ 8040 triple quadruple mass spectrometer (Shimadzu, Kyoto, Japan), and the column used Ultra-2 (25 m × 0.20 mm I.D., 0.11 μm film thickness) (Agilent Technologies, Atlanta, GA, USA)[1, 2]. Helium was used as the carrier gas at a constant flow rate of 0.5 mL/min. Argon was used as the collision gas. Samples (1.0 μL) were injected in split-injection mode (10:1). For FA and OA profiling analyses, the oven temperature was initially set at 100℃ for 2 min and increased to 300℃ at a rate of 10℃/min, with a holding time of 8 min. For AA profiling analysis, the oven temperature was initially set at 140℃ for 3 min and increased to 300℃ at a rate of 8℃/min, with a holding time of 5 min. Ionization was performed in electron impact (EI) mode at 70 eV.

##### LC-MS/MS analysis

LC-MS/MS analysis of AAs, kynurenine pathway metabolites and nucleosides was performed using a Triple Quadrupole LCMS-8050 system (Shimadzu, Kyoto, Japan) equipped with a binary pump (LC-30AD), a column oven (CTO20AC), an autosampler (SIL-30AC), and a system controller (CMB-20A). An Intrada Amino Acid (3.0 x 100 mm ID, 3 μm particle size from Seoul, Korea) column, Kinetex (4.6×250 mm ID, 5 μm particle size) column, and Synergi Hydro-RP (4.6×150 mm ID, 4 μm particle size) column from Phenomenex (Phennomenex, Macclesfeld, UK) were used for analyses of AAs, kynurenine, purine, and pyrimidine pathway-related metabolites, respectively. Solvents used for analysis of AAs were as follows: (A) acetonitrile/tetrahydrofuran/25 mM ammonium formate/formic acid = 9/75/16/0.3 (v/v/v/v) and (B) acetonitrile/100 mM ammonium formate = 20/80 (v/v). Solvents used for analysis of kynurenine pathway-related metabolites were as follows: (A) distilled water containing 0.1% formic acid and (B) acetonitrile containing 0.1% formic acid. Solvents used for analysis of purine and pyrimidine pathway-related metabolites were as follows: (A) 10 mM ammonium acetate in distilled water and (B) methanol. For AA profiling analysis, gradient elution of mobile phase B was initially set at 0% for 2.5 min, increased to 17% (6.5 min), and then increased from 17% to 100% (10 min) with a holding time of 2 min. For kynurenine pathway-related metabolite profiling analysis, mobile phase B was initially set at 0% for 6 min and then increased to 100% (15 min) with a holding time of 8 min. For purine and pyrimidine pathway-related metabolite profiling analysis, mobile phase B was initiated at 15% (10 min) with a flow rate of 0.3 mL/min, increased from 15% to 60% (1.0 min), held at 60% for 6 min, increased from 60% to 100% (0.5 min), and then held at 100% for 5.5 min. Mass spectrometric detection was operated in electrospray ionization (ESI) positive mode. Multiple reaction monitoring (MRM) mode was used for the quantification of metabolites. The nebulizing gas flow was 2.0 L/min. The drying and heating gas flow rates were both 10.0 L/min. The interface temperature was 300°C. The DL-temperature and heat block temperature were 250°C and 400°C respectively. The collision induced dissociation (CID) gas pressure was 270 kPa. The ion transitions in SRM mode were optimized according to the declustering potential (DP), entrance potential (EP) and collision energy (CE).

#### Sample preparation for profiling analyses of FAs, OAs and AAs in plasma by GC-MS/MS

Profiling analyses of FAs, OAs and AAs were performed by GC–MS/MS of TBDMS derivative, MO-TBDMS derivatives, and EOC-TBDMS derivatives, respectively, as previously described [1,2]. Briefly, proteins were removed using acetonitrile from 100 µL of serum containing pentadecanoic acid (0.1 µg) for the IS of FAs,$^{13}C_2$-succinic acid and 3,4-dimethoxybenzoic acid (0.5 and 0.1 µg) for the IS of OAs, and norvaline, $^{13}C_1$-leucine, and $^{13}C_1$-phenylalanine (0.2, 0.4, and 0.5 µg) for the ISs of AAs, respectively. After centrifugation, the supernatant was spiked into distilled water (1.0 mL), adjusted to pH < 2.0 with 10% $H_2SO_4$, saturated with sodium chloride, and sequentially extracted with diethyl ether (3 mL) and ethyl acetate (2.0 mL). The FA and OA extracts containing TEA were evaporated to dryness under a gentle stream of nitrogen (40°C). Prior to GC–MS/MS analysis, TBDMS derivatives were produced in a toluene (10 µL) and MTBSTFA (20 µL) mixture for 60 min at 60°C. The derivatives were transferred to autovials and analyzed directly by GC–MS/MS in MRM mode.

#### Sample preparation for profiling analysis of AAs, kynurenine pathway metabolites and nucleosides in plasma by LC–MS/MS

For profiling analysis of AAs, kynurenine pathway metabolites and nucleosides, proteins were removed using acetonitrile for 50 µL of plasma containing $^{13}C_1$-leucine, and $^{13}C_1$-phenylalanine (25 ng, 50 ng) as the IS of AAs, 3,4-dimethoxybenzoic acid (2.5 µg) as IS of kynurenine pathway metabolites, and 3-deazauridine (5 ng) as IS of nucleosides. After centrifugation, the supernatants were transferred to autovials and analyzed directly by LC–MS/MS in MRM mode.

#### Results

##### Metabolic profiling and univariate analyses in plasma

Considering the relatively small number of cases with NASH, we also profiled metabolites that showed a more than 20% difference in between-group comparisons with or without statistical significance.

In the NAFL group compared to the control group, six AAs (cysteine, aspartic acid, glutamic acid, tryptophan, tyrosine, and α-aminoadipic acid), two kynurenine pathway metabolites (kynurenine and kynurenic acid), three nucleosides (5,6-dihydrouridine, pseudouridine, and 1-methyladenosine), eight OAs (2-hydroxybutyric acid, 3-hydroxybutyric acid, 3-hydroxypropionic acid, α-ketoglutaric acid, malic acid, 2-hydroxyglutaric acid, *cis*-aconitic acid, and 4-hydroxyphenylactic acid), and five FAs (myristoleic acid, palmitoleic acid, γ-linolenic acid, α-linolenic acid, and 3-hydroxyoctadecanoic acid) were increased by more than 20%. However, 4-hydroxyproline was decreased by more than 20% in the NAFL group compared to the healthy control group.

In the NASH group compared to the healthy control group, four AAs (cysteine, aspartic acid, glutamic acid, tryptophan, and tyrosine), kynurenic acid, 13 OAs (pyruvic acid, acetoacetic acid, glycolic acid, 2-hydroxybutyric acid, 3-hydroxybutyric acid, 3-hydroxypropionic acid, fumaric acid, oxaloacetic acid, α-ketoglutaric acid, malic acid, 2-

hydrxoyglutaric acid, *cis*-aconitic acid, and isocitric acid), and six FAs (myristoleic acid, palmitoleic acid, γ-linolenic acid, oleic acid, α-linolenic acid, and docosatetraenoic acid) were increased, while five AAs (4-hydroxyproline, creatine, citrulline, 1-methylhistidine, and arginine) and erucic acid were decreased by more than 20%.

In the NASH group compared to the NAFL group, three AAs (aspartic acid, glutamic acid, and sarcosine), eight OAs (3-hydroxybutyric acid, fumaric acid, oxaloacetic acid, α-ketoglutaric acid, malic acid, 2-hydroxyglutaric acid, *cis*-aconitic acid, and isocitric acid), and two FAs (myristoleic acid and palmitoleic acid) were increased, while three AAs (α-aminoadipic acid, creatine, and citrulline) and erucic acid were decreased by more than 20%.

In particular, four AAs (aspartic acid, 39% to 133%; glutamic acid, 95% to 143%; tyrosine, 40% to 61%; and 4-hydroxyproline, -33% to -38%), 13 OAs (pyruvic acid, 17% to 23%; acetoacetic acid, 5% to 160%; glycolic acid, 12% to 28%; 2-hydroxybutyric acid, 38% to 62%; 3-hydroxybutyric acid, 23% to 110%; 3-hydroxypropionic acid, 38% to 54%; fumaric acid, 15% to 47%; oxaloacetic acid, 9% to 45%; α-ketoglutaric acid, 54% to 118%; malic acid, 78% to 184%; 2-hydroxyglutaric acid, 35% to 68%; *cis*-aconitic acid, 27% to 65%; and isocitric acid, 11% to 38%), and five FAs (myristoleic acid, 75% to 150%; palmitoleic acid, 69% to 120%; γ-linolenic acid, 43% to 59%; α-linolenic acid, 38% to 42%; and docosatetraenoic acid, 13% to 21%) gradually increased or decreased as NAFLD progressed from NAFL to NASH. Four AAs (creatine, 21%; citrulline, 33%; 1-methylhistidine, 30%; arginine, 21%; and erucic acid, -30%) were decreased by more than 20% in only the NASH group.

**Abbreviations**

AA, amino acid; FA, fatty acid; GC-MS/MS, gas chromatography-tandem mass spectrometry; LC-MS/MS, liquid chromatography-MS/MS; OA, organic acid; NAFLD, nonalcoholic fatty liver disease; NAFL, nonalcoholic fatty liver; NASH, nonalcoholic steatohepatitis; SD, standard deviation.

### Supplementary Tables

#### Table S1. Levels of metabolites in plasma from the control, NAFL, and NASH groups

| No. | Metabolites | Concentration (ng/μL) Mean ± SD | | | Normalized value[a] | | Wilcoxon rank-sum test P-value | | | Kruskal-Wallis Test 3 Groups | |
|---|---|---|---|---|---|---|---|---|---|---|---|
| | | Control | NAFL | NASH | NAFL | NASH | Control vs NAFL | Control vs NASH | NAFL vs NASH | P-value | Q-value[b] |
| | **Amino acids** | | | | | | | | | | |
| 1. | Alanine | 29.40 ± 8.43 | 33.52 ± 7.42 | 34.77 ± 6.07 | 1.14 | 1.18 | 0.036 | 0.016 | 0.752 | 0.039 | 0.163 |
| 2. | Glycine | 10.80 ± 3.93 | 8.86 ± 2.84 | 10.03 ± 4.75 | 0.82 | 0.93 | 0.009 | 0.102 | 0.823 | 0.036 | 0.163 |
| 3. | α-Aminobutyric acid | 2.27 ± 0.69 | 2.31 ± 0.85 | 2.36 ± 0.90 | 1.02 | 1.04 | 0.923 | 0.707 | 0.638 | 0.881 | 0.916 |
| 4. | Valine | 29.17 ± 5.36 | 33.51 ± 7.13 | 32.46 ± 5.77 | 1.15 | 1.11 | 0.012 | 0.034 | 0.638 | 0.026 | 0.149 |
| 5. | β-Aminoisobutyric acid | 0.28 ± 0.17 | 0.30 ± 0.22 | 0.27 ± 0.25 | 1.08 | 0.97 | 0.944 | 0.372 | 0.399 | 0.620 | 0.753 |
| 6. | Leucine | 17.00 ± 4.00 | 18.68 ± 4.45 | 18.46 ± 4.76 | 1.10 | 1.09 | 0.185 | 0.398 | 0.728 | 0.393 | 0.596 |
| 7. | Isoleucine | 19.22 ± 4.66 | 22.38 ± 6.55 | 21.75 ± 5.48 | 1.16 | 1.13 | 0.066 | 0.102 | 0.920 | 0.132 | 0.348 |
| 8. | Proline | 25.30 ± 6.85 | 27.74 ± 7.46 | 28.54 ± 6.73 | 1.10 | 1.13 | 0.165 | 0.137 | 0.616 | 0.236 | 0.482 |
| 9. | Pipecolic acid | 0.25 ± 0.16 | 0.23 ± 0.09 | 0.25 ± 0.11 | 0.91 | 1.00 | 0.638 | 0.526 | 0.521 | 0.713 | 0.786 |
| 10. | Pyroglutamic acid | 6.09 ± 3.29 | 6.65 ± 2.56 | 7.14 ± 2.30 | 1.09 | 1.17 | 0.281 | 0.150 | 0.435 | 0.291 | 0.500 |
| 11. | Serine | 27.58 ± 8.17 | 25.37 ± 9.15 | 29.12 ± 8.19 | 0.92 | 1.06 | 0.253 | 0.573 | 0.104 | 0.210 | 0.462 |
| 12. | Threonine | 78.56 ± 28.72 | 69.24 ± 30.32 | 70.84 ± 29.01 | 0.88 | 0.90 | 0.384 | 0.690 | 0.982 | 0.716 | 0.786 |
| 13. | Phenylalanine | 5.01 ± 1.43 | 5.84 ± 2.38 | 5.93 ± 2.58 | 1.17 | 1.19 | 0.287 | 0.268 | 0.982 | 0.462 | 0.651 |
| 14. | Cysteine | 0.49 ± 0.50 | 0.72 ± 0.75 | 0.65 ± 0.63 | 1.48 | 1.34 | 0.406 | 0.573 | 0.859 | 0.686 | 0.786 |
| 15. | Aspartic acid | 18.57 ± 9.50 | 25.86 ± 22.78 | 43.18 ± 80.63 | 1.39 | 2.33 | 0.330 | 0.007 | 0.060 | 0.031 | 0.162 |
| 16. | 4-Hydroxyproline | 30.56 ± 25.42 | 20.53 ± 12.50 | 18.85 ± 11.48 | 0.67 | 0.62 | 0.305 | 0.164 | 0.391 | 0.302 | 0.507 |
| 17. | Glutamic acid | 6.52 ± 2.47 | 12.74 ± 6.45 | 15.88 ± 6.36 | 1.95 | 2.43 | <0.001 | <0.001 | 0.011 | <0.001 | <0.001 |
| 18. | Tryptophan | 19.66 ± 15.20 | 24.01 ± 19.48 | 25.30 ± 33.96 | 1.22 | 1.29 | 0.330 | 0.656 | 0.616 | 0.590 | 0.753 |
| 19. | Tyrosine | 20.15 ± 9.59 | 28.21 ± 18.65 | 32.41 ± 19.79 | 1.40 | 1.61 | 0.003 | <0.001 | 0.261 | 0.001 | 0.015 |
| 20. | Methionine | 5.07 ± 1.29 | 4.99 ± 1.46 | 5.39 ± 1.41 | 0.98 | 1.06 | 0.872 | 0.481 | 0.341 | 0.620 | 0.753 |
| 21. | α-Aminoadipic acid | 0.54 ± 0.24 | 0.83 ± 0.44 | 0.62 ± 0.34 | 1.55 | 1.16 | 0.003 | 0.557 | 0.091 | 0.012 | 0.087 |
| 22. | Sarcosine | 2.07 ± 1.64 | 2.00 ± 0.95 | 2.41 ± 2.35 | 0.97 | 1.16 | 0.311 | 0.606 | 0.740 | 0.591 | 0.753 |
| 23. | Creatine | 26.68 ± 15.97 | 27.49 ± 14.20 | 21.19 ± 12.58 | 1.03 | 0.79 | 0.954 | 0.335 | 0.091 | 0.287 | 0.500 |
| 24. | Glutamine | 174.09 ± 68.05 | 168.51 ± 58.24 | 151.61 ± 78.46 | 0.97 | 0.87 | 0.974 | 0.359 | 0.318 | 0.553 | 0.741 |
| 25. | Creatinine | 6.26 ± 2.19 | 5.59 ± 2.68 | 5.57 ± 2.32 | 0.89 | 0.89 | 0.248 | 0.372 | 0.871 | 0.469 | 0.651 |

| | | | | | | | | | | |
|---|---|---|---|---|---|---|---|---|---|---|
| 26. Asparagine | 8.50 ± 3.57 | 8.75 ± 3.84 | 7.49 ± 3.40 | 1.03 | 0.88 | 0.832 | 0.335 | 0.195 | 0.415 | 0.619 |
| 27. Citrulline | 6.51 ± 3.92 | 6.55 ± 3.26 | 5.00 ± 1.92 | 1.01 | 0.77 | 0.782 | 0.084 | 0.080 | 0.149 | 0.381 |
| 28. 1-Methylhistidine | 0.44 ± 0.21 | 0.37 ± 0.18 | 0.31 ± 0.16 | 0.84 | 0.70 | 0.173 | 0.026 | 0.268 | 0.086 | 0.283 |
| 29. Histidine | 13.48 ± 4.04 | 12.35 ± 4.23 | 11.38 ± 3.86 | 0.92 | 0.84 | 0.264 | 0.108 | 0.463 | 0.252 | 0.482 |
| 30. 3-Methylhistidine | 2.44 ± 0.47 | 2.64 ± 1.88 | 2.15 ± 0.09 | 1.08 | 0.88 | 0.567 | 0.018 | 0.054 | 0.055 | 0.218 |
| 31. Lysine | 33.51 ± 18.73 | 36.97 ± 22.01 | 30.79 ± 19.81 | 1.10 | 0.92 | 0.638 | 0.622 | 0.303 | 0.580 | 0.753 |
| 32. Ornithine | 8.79 ± 4.76 | 9.47 ± 5.21 | 8.49 ± 4.51 | 1.08 | 0.97 | 0.812 | 0.690 | 0.616 | 0.853 | 0.910 |
| 33. Arginine | 14.19 ± 6.28 | 13.61 ± 6.54 | 11.20 ± 5.12 | 0.96 | 0.79 | 0.611 | 0.124 | 0.180 | 0.251 | 0.482 |
| **Kynurenine pathway metabolites and nucleosides** | | | | | | | | | | |
| 34. Choline | 0.97 ± 0.37 | 1.00 ± 0.37 | 1.02 ± 0.33 | 1.03 | 1.05 | 0.541 | 0.359 | 0.728 | 0.655 | 0.785 |
| 35. Kynurenine | 0.43 ± 0.13 | 0.52 ± 0.19 | 0.50 ± 0.21 | 1.21 | 1.17 | 0.061 | 0.279 | 0.705 | 0.174 | 0.406 |
| 36. Xanthurenic acid | 0.17 ± 0.05 | 0.19 ± 0.05 | 0.19 ± 0.04 | 1.09 | 1.08 | 0.275 | 0.312 | 0.823 | 0.464 | 0.651 |
| 37. Kynurenic acid | 0.005 ± 0.006 | 0.010 ± 0.009 | 0.009 ± 0.006 | 1.88 | 1.70 | <0.001 | 0.007 | 0.740 | 0.001 | 0.015 |
| 38.. 5,6-Dihydrouridine | 0.22 ± 0.15 | 0.26 ± 0.25 | 0.21 ± 0.13 | 1.20 | 0.99 | 0.403 | 0.687 | 0.649 | 0.672 | 0.786 |
| 39. Pseudouridine | 0.92 ± 0.38 | 1.22 ± 1.16 | 1.03 ± 0.57 | 1.33 | 1.12 | 0.138 | 0.477 | 0.365 | 0.275 | 0.494 |
| 40. Uridine | 1.48 ± 0.66 | 1.64 ± 1.09 | 1.39 ± 0.77 | 1.10 | 0.94 | 0.464 | 0.924 | 0.511 | 0.688 | 0.786 |
| 41. 1-Methyladenosine | 0.002 ± 0.001 | 0.002 ± 0.001 | 0.002 ± 0.001 | 1.21 | 1.02 | 0.262 | 0.653 | 0.122 | 0.243 | 0.482 |
| **Organic acids** | | | | | | | | | | |
| 42. Pyruvic acid | 9.48 ± 3.44 | 11.08 ± 4.01 | 11.69 ± 3.26 | 1.17 | 1.23 | 0.107 | 0.041 | 0.562 | 0.107 | 0.308 |
| 43. Acetoacetic acid | 21.41 ± 21.23 | 22.45 ± 17.37 | 55.61 ± 106.45 | 1.05 | 2.60 | 0.384 | 0.030 | 0.130 | 0.095 | 0.290 |
| 44. Lactic acid | 477.42 ± 167.15 | 501.71 ± 235.17 | 518.95 ± 189.41 | 1.05 | 1.09 | 0.357 | 0.137 | 0.627 | 0.366 | 0.579 |
| 45. Glycolic acid | 64.06 ± 9.99 | 71.68 ± 19.68 | 81.84 ± 37.73 | 1.12 | 1.28 | 0.132 | 0.046 | 0.417 | 0.113 | 0.308 |
| 46. 2-Hydroxybutyric acid | 4.54 ± 1.76 | 6.28 ± 3.43 | 7.34 ± 3.57 | 1.38 | 1.62 | 0.033 | 0.003 | 0.118 | 0.008 | 0.074 |
| 47. 3-Hydroxybutyric acid | 6.62 ± 12.72 | 8.17 ± 11.54 | 13.87 ± 22.45 | 1.23 | 2.10 | 0.084 | 0.012 | 0.195 | 0.033 | 0.163 |
| 48. 3-Hydroxypropionic acid | 6.88 ± 3.36 | 9.52 ± 4.27 | 10.59 ± 4.86 | 1.38 | 1.54 | 0.010 | 0.002 | 0.349 | 0.005 | 0.060 |
| 49. Malonic acid | 0.11 ± 0.07 | 0.10 ± 0.05 | 0.09 ± 0.04 | 0.86 | 0.81 | 0.872 | 0.557 | 0.752 | 0.869 | 0.916 |
| 50. Succinic acid | 9.15 ± 2.03 | 9.01 ± 3.03 | 9.93 ± 2.98 | 0.98 | 1.09 | 0.676 | 0.119 | 0.190 | 0.262 | 0.482 |
| 51. Fumaric acid | 1.06 ± 1.33 | 1.23 ± 1.55 | 1.56 ± 1.50 | 1.15 | 1.47 | 0.933 | 0.203 | 0.236 | 0.389 | 0.596 |
| 52. Oxaloacetic acid | 0.75 ± 0.44 | 0.82 ± 0.45 | 1.09 ± 0.52 | 1.09 | 1.45 | 0.350 | 0.004 | 0.025 | 0.015 | 0.101 |
| 53. a-Ketoglutaric acid | 1.70 ± 0.56 | 2.63 ± 1.32 | 3.71 ± 1.85 | 1.54 | 2.18 | 0.002 | <0.001 | 0.033 | <0.001 | 0.004 |
| 54. Malic acid | 1.25 ± 1.32 | 2.23 ± 2.71 | 3.55 ± 3.22 | 1.78 | 2.84 | 0.281 | 0.003 | 0.077 | 0.024 | 0.144 |
| 55. 2-Hydroxyglutaric acid | 1.17 ± 0.61 | 1.57 ± 1.16 | 1.96 ± 1.74 | 1.35 | 1.68 | 0.169 | 0.043 | 0.318 | 0.113 | 0.308 |
| 56. *cis*-Aconitic acid | 0.08 ± 0.04 | 0.10 ± 0.05 | 0.13 ± 0.06 | 1.27 | 1.65 | 0.051 | 0.002 | 0.151 | 0.010 | 0.082 |
| 57. 4-Hydroxyphenyllactic acid | 0.10 ± 0.06 | 0.12 ± 0.07 | 0.11 ± 0.07 | 1.21 | 1.19 | 0.475 | 0.481 | 0.932 | 0.713 | 0.786 |
| 58. Citric acid | 4.83 ± 3.34 | 4.53 ± 2.69 | 4.53 ± 2.55 | 0.94 | 0.94 | 0.964 | 0.925 | 0.969 | 1.000 | 1.000 |
| 59. Isocitric acid | 0.28 ± 0.11 | 0.31 ± 0.14 | 0.39 ± 0.11 | 1.11 | 1.38 | 0.363 | 0.003 | 0.011 | 0.009 | 0.077 |
| **Fatty acids** | | | | | | | | | | |
| 60. Myristoleic acid | 0.13 ± 0.08 | 0.22 ± 0.15 | 0.31 ± 0.19 | 1.74 | 2.50 | 0.002 | <0.001 | 0.085 | <0.001 | 0.004 |

| | | | | | | | | | | | |
|---|---|---|---|---|---|---|---|---|---|---|---|
| 61. | Palmitoleic acid | 3.26 ± 1.87 | 5.53 ± 3.97 | 7.18 ± 5.16 | 1.69 | 2.20 | 0.009 | 0.001 | 0.268 | 0.004 | 0.048 |
| 62. | Palmitic acid | 46.36 ± 14.60 | 51.46 ± 16.61 | 54.28 ± 28.11 | 1.11 | 1.17 | 0.110 | 0.248 | 0.763 | 0.247 | 0.482 |
| 63. | γ-Linolenic acid | 1.38 ± 0.96 | 1.97 ± 1.48 | 2.20 ± 1.20 | 1.43 | 1.59 | 0.177 | 0.020 | 0.357 | 0.091 | 0.286 |
| 64. | Linoleic acid | 29.52 ± 12.52 | 31.64 ± 12.48 | 34.79 ± 20.09 | 1.07 | 1.18 | 0.317 | 0.335 | 0.920 | 0.520 | 0.709 |
| 65. | Oleic acid | 35.71 ± 13.41 | 42.23 ± 17.39 | 49.51 ± 27.33 | 1.18 | 1.39 | 0.132 | 0.098 | 0.562 | 0.175 | 0.406 |
| 66. | α-Linolenic acid | 1.91 ± 0.85 | 2.65 ± 1.31 | 2.71 ± 1.15 | 1.38 | 1.42 | 0.025 | 0.022 | 0.871 | 0.037 | 0.163 |
| 67. | Octadecanoic acid | 18.41 ± 5.42 | 17.92 ± 5.18 | 17.73 ± 4.19 | 0.97 | 0.96 | 0.872 | 1.000 | 0.994 | 0.992 | 1.000 |
| 68. | Arachidonic acid | 10.40 ± 8.30 | 9.60 ± 7.32 | 9.84 ± 10.75 | 0.92 | 0.95 | 0.451 | 0.359 | 0.740 | 0.608 | 0.753 |
| 69. | 11-Eicosenic acid | 0.44 ± 0.13 | 0.48 ± 0.20 | 0.48 ± 0.17 | 1.10 | 1.10 | 0.515 | 0.589 | 0.871 | 0.772 | 0.835 |
| 70. | Eicosanoic acid | 0.16 ± 0.05 | 0.15 ± 0.04 | 0.15 ± 0.04 | 0.91 | 0.91 | 0.122 | 0.088 | 0.752 | 0.175 | 0.406 |
| 71. | Docosatetraenoic acid | 4.79 ± 1.95 | 5.43 ± 3.50 | 5.80 ± 4.25 | 1.13 | 1.21 | 0.933 | 0.815 | 0.823 | 0.962 | 0.987 |
| 72. | Docosapentaenoic acid (DPA) | 1.90 ± 0.66 | 2.19 ± 0.71 | 2.09 ± 0.80 | 1.15 | 1.10 | 0.029 | 0.108 | 0.391 | 0.063 | 0.228 |
| 73. | 2-Hydroxyoctadecanoic acid | 0.11 ± 0.02 | 0.11 ± 0.01 | 0.11 ± 0.02 | 1.00 | 1.01 | 0.620 | 0.164 | 0.230 | 0.320 | 0.517 |
| 74. | Erucic acid | 0.71 ± 0.47 | 0.63 ± 0.36 | 0.50 ± 0.12 | 0.90 | 0.70 | 0.704 | 0.049 | 0.041 | 0.078 | 0.268 |
| 75. | Docosanoic acid | 0.22 ± 0.05 | 0.20 ± 0.02 | 0.20 ± 0.01 | 0.92 | 0.90 | 0.107 | 0.171 | 0.728 | 0.210 | 0.462 |
| 76. | 3-Hydroxyoctadecanoic acid | 1.41 ± 2.55 | 1.76 ± 2.97 | 1.17 ± 2.43 | 1.24 | 0.83 | 0.638 | 0.203 | 0.349 | 0.431 | 0.630 |
| 77. | Nervonic acid | 0.40 ± 0.10 | 0.36 ± 0.08 | 0.37 ± 0.08 | 0.91 | 0.93 | 0.161 | 0.248 | 0.982 | 0.321 | 0.517 |
| 78. | Tetracosanoic acid | 0.33 ± 0.06 | 0.31 ± 0.08 | 0.32 ± 0.06 | 0.95 | 0.99 | 0.149 | 0.164 | 0.920 | 0.262 | 0.482 |
| 79. | Hexacosanoic acid | 0.41 ± 0.35 | 0.33 ± 0.08 | 0.35 ± 0.07 | 0.81 | 0.84 | 0.025 | 0.072 | 0.932 | 0.061 | 0.228 |

Levels of metabolites are expressed as the mean ± SD. Plasma metabolites were identified by targeted metabolomics analysis.

[a],Values normalized to the corresponding mean value of each metabolite in the control group.

[b], *P* values adusted by the false discovery rate.

#### Figure legends

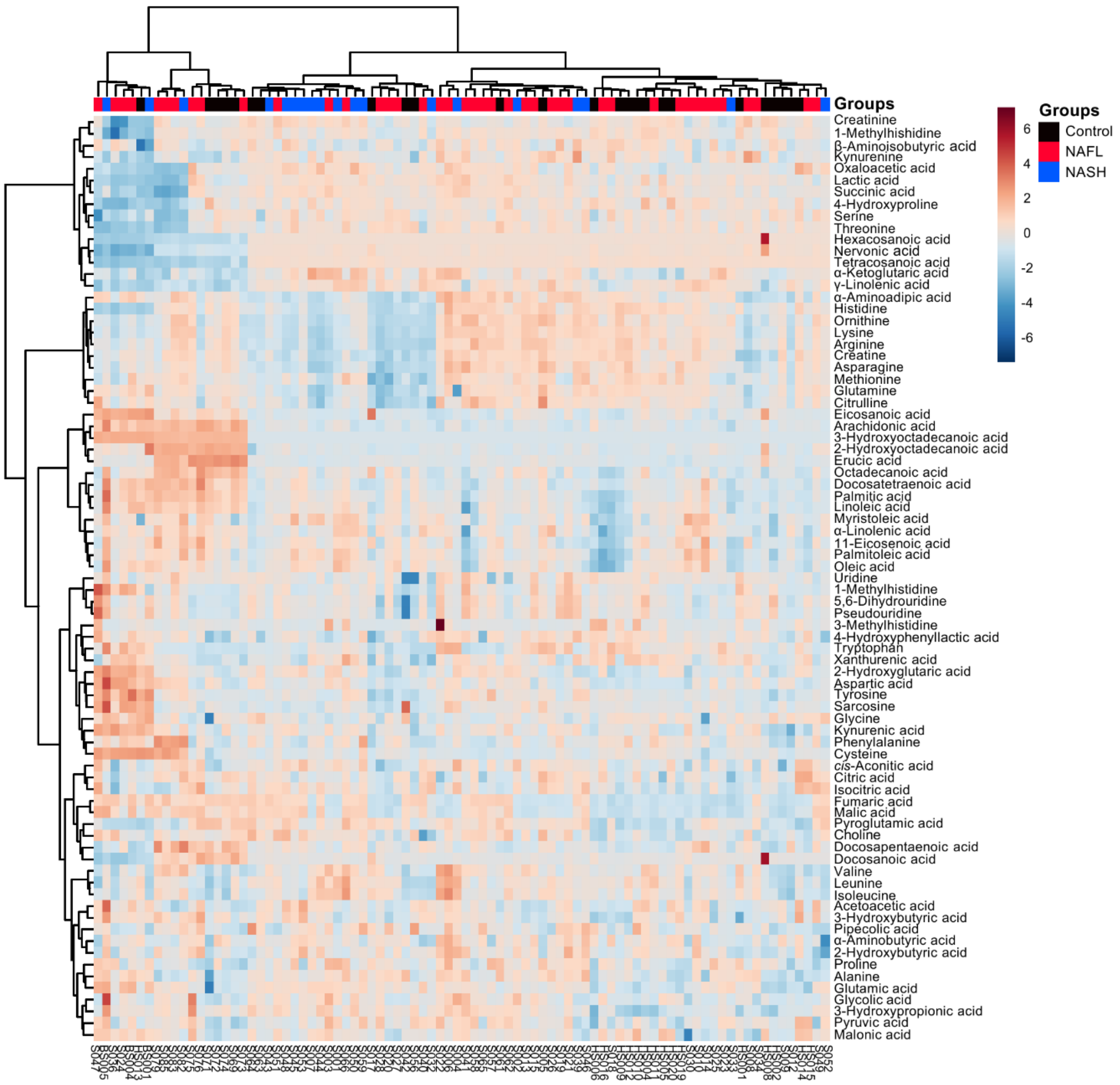


**Figure S1. Hierarchical clustering heatmaps showing the normalized plasma metabolite levels (Z scores) of all participants.**

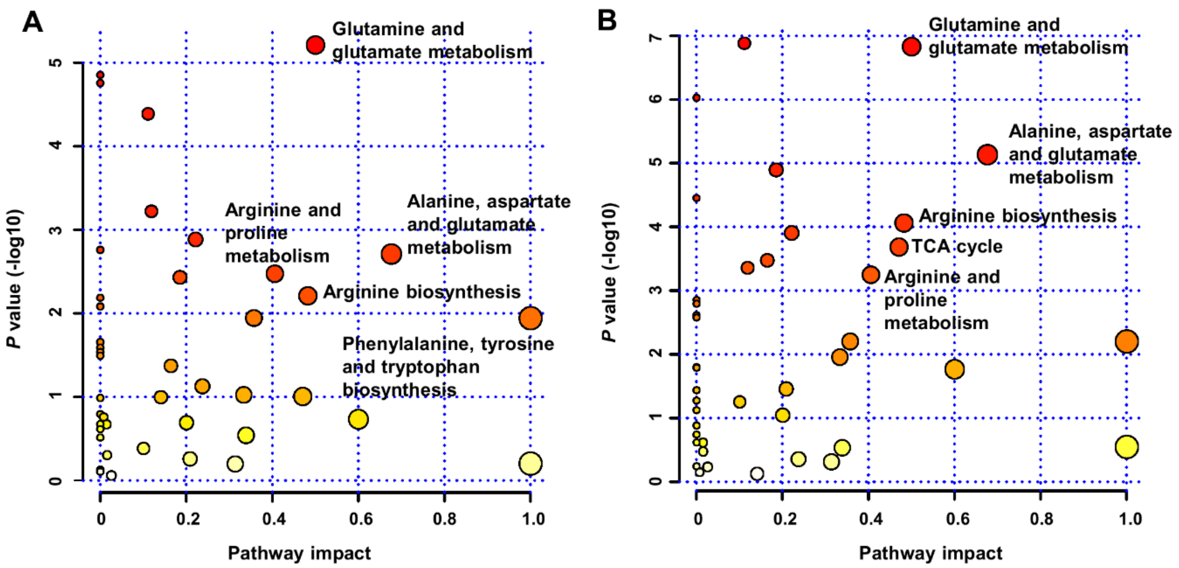


**Figure S2. Bubble plots of altered metabolic pathways related to changes in plasma metabolites in the NAFL versus control group (A) and in the NASH group versus control groups (B).**

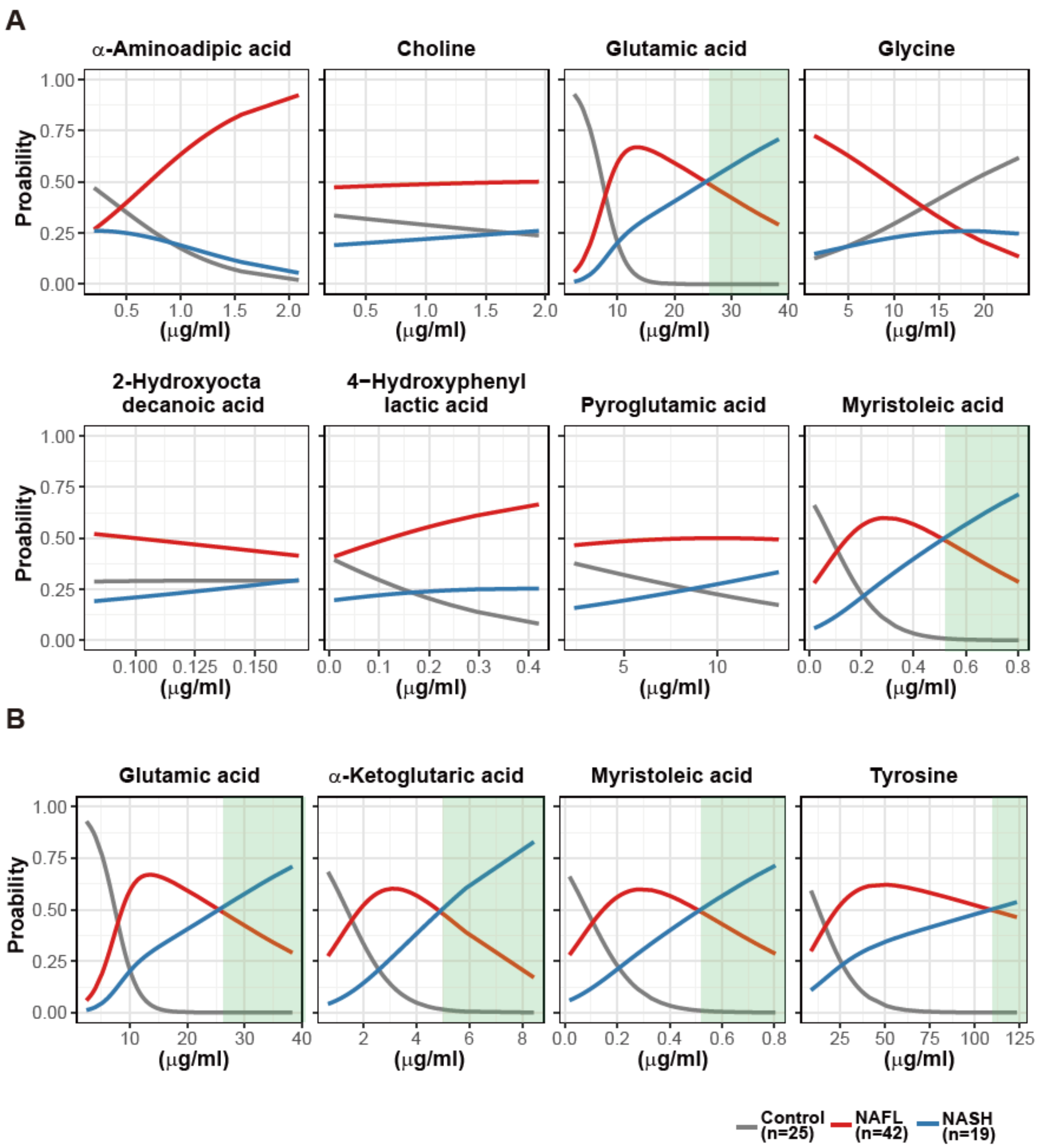


**Figure S3. Multinomial logistic regression (MLR) estimating the performance of RF-featured metabolites in between-group discrimination.** (**A**) Eight metabolites selected for discrimination between the control and NAFL groups. (**B**) Four metabolites selected for discrimination between the control and NASH groups. Green windows indicate the zone distinguishing NASH from non-NASH.
